# Retrospective Assessment of Treatments of Hospitalized Covid-19 Patients

**DOI:** 10.1101/2021.04.20.21255792

**Authors:** Ghooi Ravindra, Lele Chitra, Motwani Madhur, Dixit Santosh, Premnath Venugopalan, Salvi Sundeep, Bhat Shreepad, Chaudhari Piyush, D’Costa Pradeep, Jahagirdar Ashwini, Mane Abhay, Padbidri Vikram, Pande Chetan, Shukla Urvi

## Abstract

Infection with SARS-Cov-2 virus, is associated with significant morbidity and mortality, in addition to the economic burden it has put on the country. While waiting for a vaccine that gives adequate protection, it is necessary to understand the course of the infection and identify drugs that could reduce its impact. The results of this multicenter study involving 1035 hospitalized patients in Pune, identified diabetes, hypertension and low lymphocyte counts as predictors of mortality. There is also an indication that multiple comorbidities add to risk of severe disease and mortality. Data from metformin treated diabetics raises the possibility of considering repurposing of this drug in a larger study. It is also noted that Hydroxychloroquine, dexamethasone, azithromycin and remdesivir were associated with lower overall mortality. Diabetes and hypertension put Covid infected patients at greater risk of death, coexistence of both diseases further augment the risk, and must be aggressively treated.

## Introduction

The pandemic infection with SARS-Cov-2 virus is one of the most serious infections to have broken out in the last century. It has caused tremendous loss on a macro and micro scale leading to untold human suffering. Despite fervent research the world over, no drug has emerged as the definitive treatment for the infection.[1] A large number of anti-viral and other drugs have been repurposed for the management of clinical sequelae, but without significant success.[2] The use of convalescent plasma too, has produced varying results, [3,4] and currently the world has put all its hope in vaccines, a few of which have received regulatory clearance.[5]

The infection and case fatality rates vary in different countries,[6] and the toll in India has been much lower than expected and predicted. [7,8] Covid infection is very dynamic, infection and case fatality rates change by the day. Despite India’s large population it has a lower infection rate than US, and case fatality rate is the lowest among the top four affected countries.[9] Various factors have been ascribed to the relative resistance of Indians to SARS-Cov-2, including BCG,[10] measles-mumps-rubella vaccines,[11] cross and herd immunity[12] and genomics.[13] It is not known whether the guidelines for management of patients issued by Indian authorities and which were revised from time to time based on available evidence was one of the factors.

In addition to treatment regimens, it is also known that some comorbidities adversely affect the prognosis of COVID-19. Individuals suffering from diabetes, chronic obstructive pulmonary disease, hypertension, chronic liver disease, ischemic heart disease had a higher risk of morbidity and mortality.[14] Early in the pandemic, the Government of India imposed lockdown and social distancing, that by some reports was effective in controlling the spread amongst the vulnerable population,[15] but others suggest that community spread had taken place by the time lockdown was introduced.[16]

Apart from the anecdotal evidence, there is no data to explain which factors have actually protected Indians from this life-threatening infection. Neither is their adequate information about whether any of these factors, individually or collectively modified the response to drugs. In addition, it is not known which among existing drugs leads to best patient outcomes. To investigate the role of co-morbidities and different treatment regimens, a retrospective assessment of treatment administered to COVID-19 patients hospitalized in Pune, between June to October 2020 was planned and conducted.

## Materials Methods

All patients hospitalized with RT-PCR positive diagnosis of Covid 19, and who either were discharged or died during hospitalization were included in the study. Patients without RT PCR confirmed infections were not included. Six hospitals located within the city of Pune and its suburbs joined the study. They were the Jehangir Hospital, Sahyadri Hospital, (branches located on Karve Road, Ahmednagar Road, and Hadapsar) Symbiosis University Hospital and Research Center and Smt. Kashibai Navale Medical College and General Hospital.

Case record forms (CRF) were created for collection of the required data. The CRF collected demographic details, basic medical details, symptoms, investigations and results, treatments used and outcomes.

Records of patients were collected from medical records of hospitals. Identifiable information of patients was not recorded. Data compilation and analysis was performed using MS Excel and R.

## Ethics

As per the National Ethical Guidelines for Biomedical and Health Research Involving Human Participants (ICMR 2017), we approached the Institutional Ethics Committee of Jehangir Clinical Development Center (JCDC) for approval. All required study documents were reviewed by the IEC and approved on 14^th^ May 2020. The IEC is registered with the Department of Health Research, Ministry of Health and Family Welfare (ECR/352/MH/2013/RR-19). As per Section 4.10.2 of the ICMR Guidelines, a single EC approval was taken and the same was conveyed to participating hospitals, whose individual ECs went through the documents and agreed to the approval given by the IEC of JCDC.

Investigators did not collect any data from patients directly, the source of data was from medical records. Patients whose details were recorded had left the hospitals, either by discharge or by death. Since only anonymized retrospective data was collected, Informed Consents were not taken from patients. Waiver from the Ethics Committee had been obtained to conduct the study without ICF as per Section 5.7 of the National Ethical Guidelines for Biomedical and Health Research Involving Human Participants of ICMR 2017.

## Results

The study totally covered 1035 patients, admitted to the above-mentioned hospitals. The demographic characteristics of the patients are given in Table 1.

**Table 1.** Demographic Characteristics of Patients.

| S. No | Parameter | Characteristic | Value | %age |
| --- | --- | --- | --- | --- |
| 1 | Age | Number | 1033 |  |
|  |  | Median | 46.0 |  |
|  |  | Mean | 46.67 |  |
|  |  | Std. Dev | 17.07 |  |
|  |  | Minimum | 5 |  |
|  |  | Maximum | 97 |  |
| 2 | Gender | Number | 1035 |  |
|  |  | Male | 654 | 63.19% |
|  |  | Female | 381 | 36.81% |
| 3 | Pregnancy | Pregnant | 10 |  |
|  |  | Non pregnant | 371 |  |
| 4 | Smoking | Smokers | 2 |  |
|  |  | Non-Smokers | 1033 |  |

In the whole cohort a total of 620 comorbidities were recorded (some patients had more than one comorbidity), the breakup of which is given in Table 2. Only two comorbidities were seen in a significant number of patients, namely diabetes and hypertension. Therefore, focus was on patients who had diabetes or hypertension, and those who had both the comorbidities simultaneously. Other comorbidities such as COPD and asthma could certainly be important in raising the risk of death, but the number of patients with these was too small to give us any meaningful inference.

**Table 2.** Comorbidities in the Cohort.

| Condition | Count | % age |
| --- | --- | --- |
| Hypertension | 297 | 28.70 |
| Diabetes | 270 | 26.09 |
| Diabetes and Hypertension | 169 | 16.33 |
| Coronary Artery Disease | 12 | 1.16 |
| Asthma | 21 | 2.03 |
| Chronic Obstructive Pulmonary Disease | 5 | 0.48 |
| Cancer | 3 | 0.29 |
| Cirrhosis | 2 | 0.19 |
| Chronic Kidney Disease | 9 | 0.87 |
| HIV | 1 | 0.10 |

Of the 1035 patients admitted, 955 (92.27%) recovered and were discharged while 80 (7.73%) patients died. The break-up of the duration of stay of these patients is given in Table 3. Patients were divided into two groups as per age, those below 46 and those above 46 years, since the median age of our cohort was 46 years.

**Table 3.** Subject Disposition by Gender and Age.

|  |  | Discharged |  | Died |  |
| --- | --- | --- | --- | --- | --- |
|  | n | n | % age | n | %age |
| All Patients | 1035 | 955 | 92.27 | 80 | 7.73 |
| Males | 654 | 604 | 92.35 | 50 | 7.65 |
| Females | 381 | 351 | 92.12 | 30 | 7.88 |
| Age < 46 | 510 | 502 | 98.43 | 8 | 1.57 |
| Age ≥ 46 | 525 | 453 | 86.28 | 72 | 13.72 |

**Table 4.** Duration of Hospitalization.

|  | Discharged<br>Mean (SD) | Died<br>Mean (SD) |
| --- | --- | --- |
| Time from Report to Outcome | 11.01 (3.99) | 10.74 (7.11) |
| Time from Admission to Outcome | 10.38 (3.94) | 11.04 (7.16) |

**Table 5.** Risk of Hypertension.

|  | n | Discharged |  | Died |  |
| --- | --- | --- | --- | --- | --- |
|  |  | n | %age | n | %age |
| Hypertensive | 297 | 246 | 82.8 | 51 | 17.1 |
| Non-Hypertensive | 738 | 709 | 96.1 | 29 | 3.9 |

Odds Ratio (OR) = 5.07, 95% Confidence Interval (CI) = (3.14, 8.18), $p < 0.0001$

**Table 6.** Risk of Diabetes.

|  | n | Discharged |  | Died |  |
| --- | --- | --- | --- | --- | --- |
|  |  | n | %age | n | %age |
| Diabetic | 270 | 223 | 82.59 | 47 | 17.41 |
| Non-Diabetic | 765 | 732 | 95.68 | 33 | 4.32 |

OR = 4.68, 95% CI = (2.92, 7.48), $p\text{-value} < 0.0001$
Note: Odds ratio corresponding to hypertension and diabetes continues to be highly significant after adjustment for age and gender.

**Table 7.** Risk of Hypertension and Diabetes.

|  | n | Discharged |  | Died |  |
| --- | --- | --- | --- | --- | --- |
|  |  | n | %age | n | %age |
| Diabetes only | 101 | 90 | 89.1 | 11 | 10.9 |
| Hypertension only | 128 | 113 | 88.3 | 15 | 11.7 |
| Diabetes & Hypertension | 169 | 133 | 78.7 | 36 | 21.3 |
| Neither comorbidity | 637 | 609 | 95.6 | 28 | 4.4 |
| Total | 1035 | 955 | 92.3 | 80 | 7.7 |

**Table 8.** Metformin and Risk of Death.

|  | n | Discharged |  | Died |  |
| --- | --- | --- | --- | --- | --- |
|  |  | n | %age | n | %age |
| Diabetes | 366 | 304 | 83.1 | 62 | 16.9 |
| Metformin | 53 | 48 | 90.6 | 5 | 9.4 |
| No Metformin | 313 | 256 | 81.8 | 57 | 18.2 |

Odds ratio for metformin usage among diabetics, OR = 0.66, CI = (0.24, 1.79), p = 0.42 The cell frequencies do not allow further analysis, for example, by gender.

**Table 9.** Role of Ventilatory Support.

|  | Enrolled | Discharged |  | Died |  |
| --- | --- | --- | --- | --- | --- |
|  | n | n | %age | n | %age |
| Total | 1035 | 955 | 92.27 | 80 | 7.73 |
| Supplemental Oxygen | 261 | 192 | 73.56 | 69 | 26.44 |
| No Supplemental Oxygen | 774 | 763 | 98.58 | 11 | 1.42 |
| Mechanical Ventilation | 92 | 24 | 26.09 | 68 | 73.91 |
| No Mechanical Ventilation | 943 | 931 | 98.73 | 12 | 1.27 |
| Non-invasive Ventilation | 72 | 24 | 33.33 | 48 | 66.67 |
| No Non-invasive Ventilation | 963 | 931 | 96.68 | 32 | 3.32 |
| Invasive Ventilation | 52 | 6 | 11.54 | 46 | 88.46 |
| No Invasive Ventilation | 983 | 949 | 96.54 | 34 | 3.46 |

Supplemental Oxygen usage: OR = 24.93, CI = (12.94, 48.02), p < 0.0001
Mechanical Ventilation usage: OR = 219.82, CI = (105.36, 458.62), p < 0.0001

**Table 10.** Severity of Infection and Mortality *.

|  | Mild |  | Moderate |  | Severe |  |
| --- | --- | --- | --- | --- | --- | --- |
| N | 787 |  | 141 |  | 107 |  |
|  | n | %age | n | %age | n | %age |
| Died | 19 | 2.41 | 23 | 16.31 | 38 | 35.51 |
| Discharge | 768 | 97.59 | 118 | 83.69 | 69 | 64.49 |

\*There was proportionally higher number of diabetics in mild, moderate, and severe infections, hence diabetes was a confounding factor.

**Table 11.** Lymphocyte Count and Risk.

|  | n | Discharged |  | Died |  |
| --- | --- | --- | --- | --- | --- |
|  |  | n | %age | n | %age |
| < 10 % | 101 | 64 | 63.3 | 37 | 36.7 |
| ≥ 10 % | 890 | 851 | 95.6 | 39 | 4.4 |
| < 15 % | 208 | 152 | 73.0 | 56 | 27.0 |
| ≥ 15 % | 783 | 763 | 97.4 | 20 | 2.6 |
| < 20 % | 321 | 257 | 80.0 | 64 | 20.0 |
| ≥ 20 % | 670 | 658 | 98.2 | 12 | 1.8 |

A trend was noted in terms of lower lymphocyte counts being associated with higher mortality. For example, for the categorization of lymphocyte count (%) as <10%, 10-20% and >20%, the Cochran-Armitage Trend test had p-value < 0.0001.

Note: Lymphocyte data was captured in percentages.

**Fig. 1.**
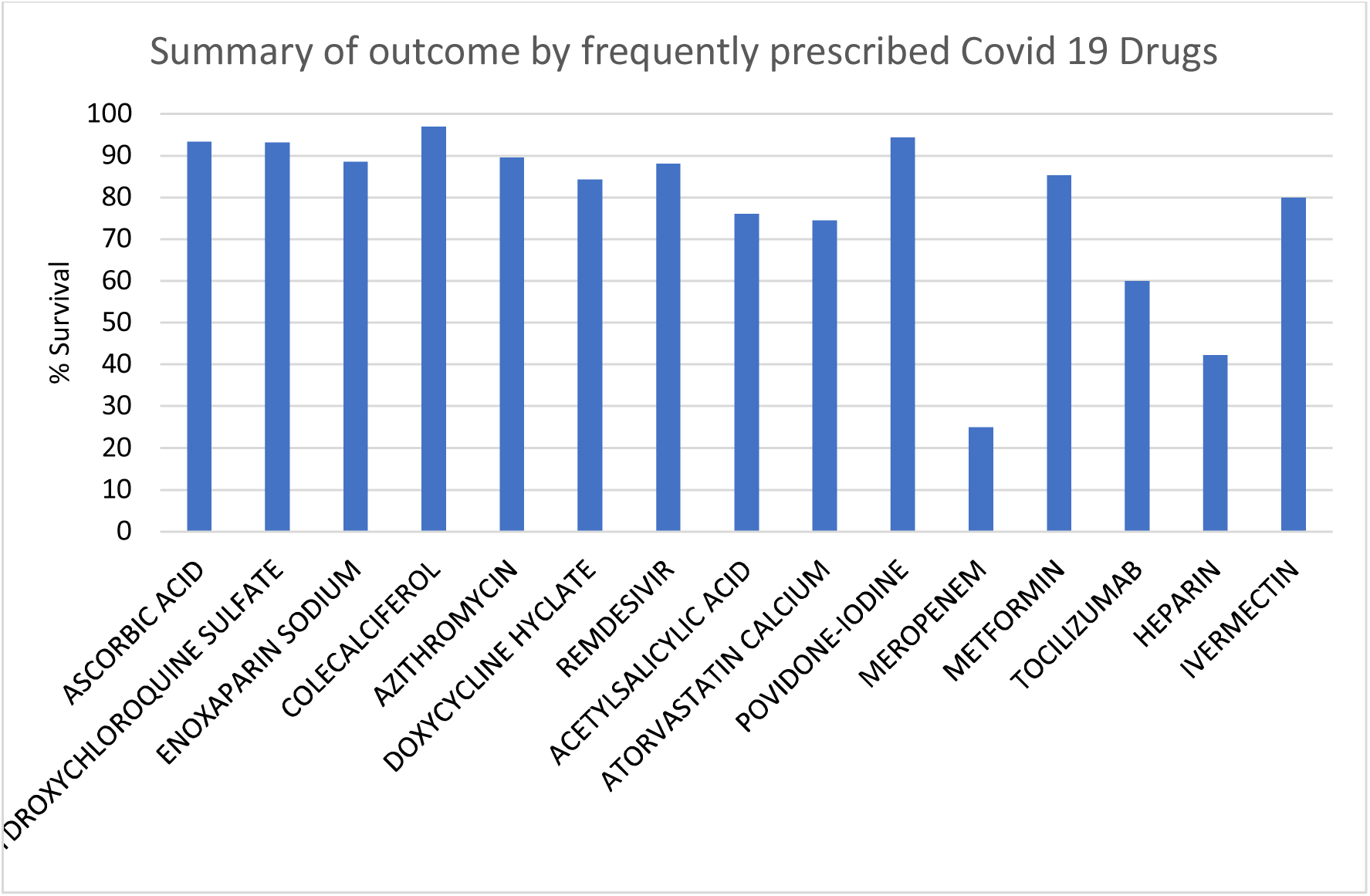
Drug Treatment and Outcome of Patients. Most patients received multiple drugs, as per the prevailing guidelines at that time. The outcome of patients receiving different drugs is shown in Figure 1. Ascorbic acid, hydroxychloroquine, cholecalciferol, and povidone iodine usage were associated with over 90% survival. Enoxaparin, azithromycin, remdesivir, metformin and ivermectin were associated with 80% to 90% survival.

## Discussion

The current study covered over 1000 patients admitted to dedicated Covid hospitals in the city of Pune from June to October 2020. Overall, 1035 patients’ data was collated, and it was found that of these 955 (92.27%) patients recovered and were discharged while 80 (7.73%) died. The death rate in our cohort, during initial hospitalization was much lower than that reported elsewhere, [17,18] we admit that patients were not followed up after discharge to record post Covid complications if any. Gender wise and age wise distribution of cases and outcome was unremarkable.

The members of the cohort were mainly hospital workers and employees. During the study period all employees that tested positive by RT PCR were immediately hospitalized on receipt of the report. Hence the duration from report to discharge or admission to discharge are very similar. So also, there was no significant difference between duration from report to death and admission to death.

Hypertension (HTN) as a comorbidity was most common followed by diabetes followed by combination of HTN and diabetes. In this series, the type of HTN (primary or secondary) was not differentiated, but HTN did increase the risk of death by around 4 times. Figures for diabetes were similar. The increased risk of both infection and mortality due to hypertension and diabetes has been widely reported. [19,20]

While diabetes is well accepted as a risk factor both for infection and poor prognosis,[21] hypertension is reported to be an important comorbidity with poor outcome.[22] Two points are however not clear, firstly, what is the risk of death in patients who suffer from both diabetes and hypertension, and secondly does the risk due to hypertension or diabetes depend upon the quality of control? The current study appears to answer the first question; however, the numbers were not adequate to estimate the odds while adjusting for the available covariates. However, the study does not answer the second question, neither was it designed to do so in the first place. It is not clear whether the risk is similar with hypertension or diabetes that is well controlled versus not well controlled. However, aggressive treatment of diabetes and hypertension in Covid patients is strongly suggested. Special care needs to be exercised when the patient has both diabetes and hypertension.

There are reports that Metformin could be used for the treatment of Covid 19, due to its immunomodulatory effects on mTOR inhibition.[23] In the current study Metformin was not used for the treatment of Covid, but to control blood sugar in a few diabetic patients. The numbers in the Metformin and non-Metformin group were not adequate to draw any definite conclusions, but it appears that those on metformin had a lower death rate than those on other drugs.

Ventilatory support is needed by a limited number of patients whose oxygen tension drops in its absence. In other words, oxygenation support is required for more serious patients, so obviously their outcomes are worse than those who do not require it. That the lung is a target for the SARS-Cov-2 virus is well known, and the use of oxygen as a rescue therapy is essential and it saves lives too.

It is axiomatic that patients with more serious clinical condition will have a poorer outcome, and this was clearly observed. Death rate increased from 2.4% to 16.31% to 35.51% in patients with mild, moderate and sever disease. The prevalence of hypertension and diabetes is also proportionately higher, hence the presence of these two co-morbidities are confounding factors while studying association between seriousness of infection and negative outcome.

Lymphocyte counts are among the factors identified by other workers as prognostic and predictive parameters.[24] There was a definite relationship between lymphocyte count and risk of death. Additionally, neutrophil/lymphocyte ratio and peak platelet/lymphocyte ratio may also have prognostic value in determining severe cases.[25]

One of the main aims of this study was to study the association of the drugs used in the management with the outcomes. In a pandemic of this sort, the first aim is to save lives, and not to evaluate drugs. As a result, very few trials have been designed to evaluate single drugs, since the treatment of patients is multi-modal. The current study too only aimed to describe associations between drugs and prognosis, given that most drugs were used as add-on to recommended treatments.

The best outcomes were associated with hydroxychloroquine, azithromycin, enoxaparin, remdesivir, doxycycline, ivermectin and tocilizumab in that order. The efficacy of each of these agents has been reported variously. [26,27] However, this was a retrospective observational study, and it cannot demonstrate the efficacy of each drug, neither was the study intended for this purpose. All that was expected of the study was to reveal the treatment that is associated with the best outcome. More studies differently designed will be required to reveal the efficacy of individual treatments identified herein.

## Data Availability

The data reported in the manuscript will be accessible with prior knowledge of the Ethics Committee overseeing the study .

## Acknowledgments

The authors would like to acknowledge the financial grant from Persistent Systems, for the conduct of this study. The support of the Clinical Team of Jehangir Clinical Development Center in data collection is gratefully acknowledged. The authors would like to thank Ms. Manali Pendse who did the data analysis.

